# Determining the cutoff points of the 5C scale for assessment of COVID-19 vaccines psychological antecedents among the Arab population: A multinational study

**DOI:** 10.1101/2021.03.27.21254461

**Authors:** Ramy Mohamed Ghazy, Samar Abd ElHafeez, Ramy Shaaban, Iffat Elbarazi, Khalid A. Kheirallah, Marwa Shawky Abdou, Ahmed Ramadan

**Affiliations:** Tropical Health Department, High Institute of Public Health, Alexandria University, Egypt; Epidemiology Department, High Institute of Public Health, Alexandria University, Egypt; Department of Instructional Technology and Learning Sciences, Utah State University, USA; Institute of Public Health, College of Medicine and Health Sciences, United Arab Emirates University, AlAin, UAE; Department of Public Health, Medical School of Jordan University of Science and Technology, Irbid 22110, Jordan; Department of demography and biostatics, Faculty of postgraduate studies for statistical research, Cairo University

**Keywords:** Arab, COVID-19, vaccine hesitancy, 5C, AUC, ROC

## Abstract

**Background:** One of the newly faced challenges during the COVID-19 is vaccine hesitancy (VH). The validated 5C scale, that assesses five psychological antecedents of vaccination, could be effective in exploring COVID-19 VH. This study aimed to determine a statistically valid cutoff points for the 5C sub-scales among the Arab population.

**Methods:** A cross-sectional study was conducted among 446 subjects from three Arab countries (Egypt, United Arab Emirates UAE, and Jordan). Information regarding sociodemographics, clinical history, COVID-19 infection and vaccination history, and 5C scale were collected online. The 5C scores were analyzed to define the cutoff points using the receiver operating characteristic curve (ROC) and to verify the capability of the questionnaire to differentiate whether responders are hesitant or non-hesitant to accept vaccination. ROC curve analysis was conducted setting for previous vaccine administration as a response, with the predictors being the main five domains of the 5C questionnaire. The mean score of each sub-scale was compared with COVID-19 vaccine intake

**Results:** The mean age of the studied population was 37±11, 42.9% were males, 44.8% from Egypt, 21.1% from Jordan, and 33.6% from UAE. Statistically significant differences between vaccinated and unvaccinated participants, respectively, weredetetd in the median score of confidence [6.0(1.3) versus 4.7(2.0)], complacency [(2.7(2.0) versus 3.0(2.0), constraints [1.7(1.7) versus 3.7(2.3)], and collective responsibility [6.7(1.7) versus 5.7(1.7)]. The area under the curve of the five scales was 0.72, 0.60, 0.76, 0.66, 0.66 for confidence, complacency, constraints, calculation, and collective responsibility at cutoff values of 5.7, 4.7, 6.0, 6.3, and 6.2, respectively.

**Conclusion:** the Arabic validated version of the 5C scale has a good discriminatory power to predict COVID-19 vaccines antecedent.

## Introduction

More than 120 million people have been affected by the 2019 coronavirus disease (COVID-19), with about 2.6 million worldwide deaths.(1) Disease containment measures should be continued until herd immunity is achieved and COVID-19 can no longer circulate. Herd immunity is acquired if at least 60% of the population are vaccinated or get infected. As an emergency response to contain the pandemic and its related burden(s), non-pharmaceutical intervention (NPI) measures were globally implemented. Thses included physical distancing and curfews. (2, 3) Mass vaccination could be the only way to build herd immunity and reduce disease burden(s).(4) With the recent introduction of several effective vaccinations, many countries scheduled mass immunization programs utilizing most emergency-approved vaccines. (4, 5)

Currently, about 13 vaccines have been approved and licensed for general use. Till now, BioNTech (Pfizer), Modern, AstraZeneca (Oxford), Sputnik (Russa), and Sinopharm/ Sinovac (China) are the most widely used vaccine.(6) Many Arab countries caught up with the global community importing and rolling out vaccines among its populations. The United Arab of Emirates (UAE), in the Arabian Gulf, was among the first countries to roll out COVID-19 vaccines with priority for the elderly, frontline workers, and patients with chronic diseases. Also, Egypt and Jordan have begun COVID-19 vaccination among health care workers and launched a governmental website for vaccine registration. On March 26, 2021, more than 78% of the targeted population living in Emirates received at least a single dose of COVID-19 vaccine, while more 3% thousand of Jordanians received COVID-19 vaccine. (7) The difference in percentages for vaccination uptake in the three aforementioned countries could be due to the time difference in starting vaccination campaigns but, most importantly, due to hesitancy toward vaccination. Vaccine hesitancy (VH) and reluctance among people in these countries and around the word is incited by lots of misinformation and dis-information circulating about vaccine side effects, lack of trust, and conspiracy theories. (8) Accordingly, exploring the prevalence of VH and its determinants is crucial using culturally adapted tools that can distinguish between hesitant and non-hesitant populations.

COVID-19 VH is part of a long-standing debate about vaccination dynamics. Vaccine hesitancy was declared by WHO as a global health threat even before the pandemic. (9) With the emergence of the COVID-19 pandemic, VH is taking a toll and seems to persist as a global public health problem that is spreading worldwide. (10). VH simply refers to a person’s refusal, or hesitation, to be vaccinated or to have their children vaccinated against a disease, even though the vaccine has been shown to be safe and efficient. (11) VH raises concerns at the individual and community levels as exposure to an infectious disease increases the risk of disease transmission among the community where effective vaccination programs are not established. (12) COVID-19 infodemic has impacted people’s perceptions and trust with offered vaccines leading to an increase in COVID-19 vaccine hesitancy even among different groups who were never known to be hesitant toward any vaccination. Moreover, intention to vaccinate is argued to be higher than the actual vaccination uptake in some countries, hence, it is essential to identify risks, intentions, and antecedents to vaccination. (13)

Betsch and colleagues recently proposed a model that synthesizes and expands prior models of VH and trust by measuring five psychological antecedents of vaccination.(14, 15) An antecedent is a psychological cause or determinant inside an individual (for example, a parent) that influences whether or not a person gets vaccinated. Confidence, complacency, constraints, calculation, and collective responsibility are the five sub-scales of this tool. The 5C scale evaluates five psychological antecedents to vaccination and offers insight into human mental representations, attitudes, and behavioral patterns that are influenced by the respondent’s environment and background.(16) While the 5C tool has been validated in different population groups, it is still not tested among Arabs. Evidence from few Arabic countries demonstrated the existence of COVID-19 vaccine hesitancy.(17) With the emergence of the COVID-19 pandemic and the distribution and manufacturing of the COVID-19 vaccine, the need to explore the existing resistance and potential hesitancy utilizing validated tools seems eminent. In this research, we aimed at identifying the cutoff point for each of the 5C sub-scale to be used in determining the psychological antecedent of the COVID-19 vaccine among the Arab population.

## Methods

### A study design and population

The data for this study were derived from a preexisting cross-sectional study conducted from January to March, 2012 to assess the prevalence of psychological antecedents for COVID-19 vaccines among the general population in the Arab world. Based on the assumptions of AUC =0.5 for the null hypothesis and AUC=0.6 for the alternative hypothesis, allocation ratio between vaccinated and non-vaccinated population was estimated to be 1 to 2, power= 0.80, and alpha error= 0.05, the minimum sample size required was estimated at 219 participants using MedCalc software application (version 19.6.3). We duplicated the sample size to allow for the comparative analysis and to compensate potential missing data.

### Data collection tools

The questionnaire was composed of three sections. The first section collects data on sociodemographics: age, sex, education, marital status, occupation (health care worker or not), and comorbidities (asthma, diabetes mellitus, hypertension, ischemic heart diseases). The second section included questions on COVID-19 infection: history of previous COVID-19 infection, family history of COVID-19 infection, mortality among relatives due to COVID-19, history of vaccination against influenza, knowledge about different types of COVID-19 vaccine, and searching the web for information about COVID19 vaccine. The third section is the validated Arabic 5C questionnaire, which is composed of 15 questions covering five main domains or subscales: confidence (Q1-Q3), complacency (Q4-Q6), constraints (Q7-Q9), calculation (Q10-Q12), and collective responsibility (Q13-Q15).

### Statistical analysis

The results are presented as means and standard deviations (SD) in case of normally distributed data, median and interquartile range (IQR) for non□normally distributed data, or as percentages for categorical data. Continuous variables were compared using t-test or Mann-Whitney test while chi-square was used to compare categorical variables.

The cutoff points for the different subscales of the Arabic version of the 5C questionnaire were determined using the receiver operating characteristic (ROC) curve based on the self-report of receiving COVID-19 vaccines. Responses to the question “HISTORY OF COVID-19 VACCINE INTAKE” was coded as a binary variable (“not vaccinated” and “vaccinated”), to distinguish between individuals who received and those who did not receive the vaccine. The ROC curve is a graphical plot of sensitivity against 1 − specificity at various discrimination cutoff points. The best cutoff point is the one that represents the best compromise between sensitivity and specificity. It will be identified using the Youden index

### Ethical consideration

The study was approved by the Ethics Committee of the Faculty of Medicine-Alexandria University/ Egypt (IRB No:00012098). All research activities were in acoordance with the International Ethical Guidelines for Epidemiological studies (18). Participants were consented online before taking the survey. Participation was voluntary and clear research objectives were presented prior to the survey.

## Results

### Participants’ Backgroun Characteristics

A total of 460 participants participated in the study. Of which, 14 were excluded due to missing data (more than 20%). Of the total 446 participants included in the analysis (Table 1) 27.6% reported a history of COVID-19 vaccine intake. The mean± SD was 37±11, 42.9% were males, 67% were married, 44.8% from Egypt, 21.1% from Jordan, and 33.6% from UAE, 40.3% had a university degree, and 35.4% were healthcare workers. More than half (63%) of participants reported a history of chronic comorbidities, 27.8% got infected with COVID-19, 35.2% gave family history of COVID-19 infection, and 21.8% had influenza vaccine, 72.9% were aware of availability of several vaccines, 10.8% reported a history of friends or relatives being infected with COVID-19 after being vaccinated, and 29.8% searched the web for information on the available vaccines.

**Table 1:** Participants’ demographics and clinical characteristics.

| Sociodemographic criteria | Total<br>(n=446) | Vaccinated<br>(n=119) | Unvaccinated<br>(n=327) | P |
| --- | --- | --- | --- | --- |
| Gender |  |  |  | <0.001 |
| Male | 191 (42.9) | 43 (36.0) | 148 (45.3) |  |
| Female | 225 (57.1) | 76 (64.0) | 179 (54.7) |  |
| Age, mean ± SD | 36.76±10.84 | 36.63±10.41 | 36.81±11.00 | 0.89 |
| Country |  |  |  | <0.001 |
| Egypt | 200 (44.8) | 15 (7.5) | 185 (92.5) |  |
| Jordan | 94 (21.1) | 7 (7.2) | 87 (92.3) |  |
| UAE | 150 (33.6) | 97 (64.7) | 53 (35.3) |  |
| Education |  |  |  | <0.001 |
| Pre university | 21 (4.7) | 12 (10.1) | 9 (2.8) |  |
| Technical/vocational | 23 (5.2) | 5 (4.2) | 18 (5.5) |  |
| University degree | 179 (40.13) | 52 (43.7) | 127 (39) |  |
| Postgraduate degree | 169 (37.9) | 49 (41.2) | 120 (36.7%) |  |
| Others | 54 (12.1) | 1 (0.8) | 53 (16.2) |  |
| Marital status |  |  |  | 0.07 |
| Single | 101(22.7) | 31 (26.1) | 70 (21.4) |  |
| Married | 301(67.5) | 82 (68.9) | 219 (67.0) |  |
| Divorced | 28(6.3) | 6 (5.0) | 22 (6.7) |  |
| Widow | 16(3.6) | 0 (0) | 16 (4.9) |  |
| Healthcare workers | 158(35.4) | 58 (48.7) | 90 (27.5) | <0.001 |
| Chronic comorbidities | 282(63%) | 31 (26.1) | 251(76.8) | <0.001 |
| Responders getting yearly Influenza vaccination | 97(21.8) | 17 (41.5%) | 80 (24.5%) | 0.02 |
| History of COVID-19 infection |  |  |  | 0.03 |
| Yes | 123(27.6) | 25 (21) | 98 (30) |  |
| No | 159(60.3) | 84 (70.6) | 185 (56.6) |  |
| Do not know | 54(12.1) | 10 (8.4) | 44 (13.5) |  |
| Family history of COVID-19 infection |  |  |  | 0.27 |
| Yes | 157(35.2) | 59 (49.6) | 139 (42.5) |  |
| No | 240(53.8) | 55 (46.2) | 161 (49.2) |  |
| Do not know | 49(11.0) | 5 (4.2) | 27 (8.3) |  |
| Family history of death due to COVID-19 infection |  |  |  | <0.001 |
| Yes | 143(32.1) | 31 (26.1) | 112 (34.3) |  |
| No | 252(56.5) | 85 (71.4) | 167 (51.1) |  |
| Do not know | 51(11.3) | 3 (2.5) | 48 (14.7) |  |
| knowledge about the availability of different vaccines ( yes ) | 325(72.9) | 112 (94.1) | 213 (65.1) | <0.001 |
| History of COVID-19 infection after vaccination among family and friends (yes ) | 48(10.8) | 40 (33.9) | 8 (8.2) | <0.001 |
| knowledge about the indications and contraindications for different types of COVID-19 vaccines | 123(27.6) | 97 (82.2) | 26 (26.8) | <0.001 |
| knowledge about the different information related to COVID-19 vaccine through searching the web | 133(29.8) | 83 (70.3) | 50 (51.5) | <0.001 |

Participants who reported receiving the COVID-19 vaccine were mainly females (57.1% vs 42.9%), aged 36.63±10.41, from UAE (64.7%), had achieved higher education (41.2% vs 36.7%), healthcare workers (48.7% vs 27.5%), did not report a history of chronic conditions (73.9 vs 23.2%), reported a history of receiving influenza vaccine yearly (41.5% vs 24.5%), did not contract COVID-19 infection (70.3% vs 56.6%), did not give a family history of COVID-19 related deaths (71.4% vs 51.1%), knew about the presence of different types of COVID-19 vaccine (82.2% vs 26.8%), and searched the web for more information about COVID-19 (70.3% vs 51.5%).

### The Arabic version of the 5C

The median and IQR were calculated for each question and 5C sub-scale. The estimated median (IQR) values for confidence (5.0(2.0)), complacency (3.0(2.0)), constrains (3.0(2.67)), calculation (6.0(1.33)), and collective responsibility (5.66(1.67)) are presented in table Table 2. The median score of each items of the 5C scale was significantly different between vaccinated and unvaccinated except for questions 5 and 12. There was a statistically significant difference between vaccinated and unvaccinated, respectively, in the median scores of confidence [6.0(1.3) versus 4.7(2.0)], complacency [(2.7(2.0) versus 3.0(2.0), constraints [1.7(1.7) versus 3.7(2.3)], and collective responsibility [6.7(1.7) versus 5.7(1.7)] in between vaccinated and unvaccinated.

**Table 2:** Median and IQR of the 5C subscales in Arabic version.

| **The subscale** | **Total** | **Vaccinated** | **Unvaccinated** | **P-value** |
| --- | --- | --- | --- | --- |
| **Median (IQR)** |  | (n=119) | (n=327) |  |
| **Confidence** | 5.0(2.0) | 6.0(1.3) | 4.66 (1.7) | **<0.001** |
| Q1: I am completely confident that vaccines are safe | 5.0(2.0) | 6.0 (3.0) | 5.0 (2.0) |  |
| Q2: Vaccinations are effective | 5.0(2.0) | 6.0 (2.0) | 5.0 (2.0) |  |
| Q3: Regarding vaccines, I am confident that public authorities decide in the best interest of the community | 5.0(2.0) | 7.0 (1.0) | 5.0 (3.0) |  |
| **Complacency** | 3.0(2.0) | 2.7 (2.0) | 3.0 (2.0) | **P<0.001** |
| Q4: Vaccination is unnecessary because vaccine-preventable diseases are not common anymore | 2.0(3.0) | 1.0 (2.0) | 3.0 (3.0) |  |
| Q5: My immune system is so strong; it also protects me against diseases | 3.0(3.0) | 4.0 (3.0) | 3.0 (3.0) |  |
| Q6: Vaccine-preventable diseases are not so severe that I should be vaccinated | 3.0(3.0) | 1.0 (2.0) | 3.0 (3.0) |  |
| **Constrains** | 3.0(2.67) | 1.7 (1.7) | 3.7 (2.3) | **P<0.001** |
| Q7: Everyday stress prevents me from being vaccinated | 3.0(3.0) | 2.0 (2.0) | 3.0 (3.0) |  |
| Q8: For me, it is inconvenient to be vaccinated | 3.0(4.0) | 1.0 (1.0) | 4.0 (3.0) |  |
| Q9: Visiting the doctor makes me feel uncomfortable; this keeps me from being vaccinated | 3.0(3.0) | 1.0 (1.0) | 3.0 (3.0) |  |
| **Calculation** | 6.0(1.33) | 6.00 (2.0) | 6.00 (1.3) | **P=0.321** |
| Q10: When I think about being vaccinated, I weigh its benefits and risks to make the best decision possible | 6.0(2.0) | 6.0 (3.0) | 6.0 (2.0) |  |
| Q11: For each and every vaccination, I closely consider whether it is useful for me. | 6.0(2.0) | 6.0 (2.0) | 6.0 (2.0) |  |
| Q12: It is important for me to fully understand the topic of vaccination before I get vaccinated | 7.0(1.25) | 7.0 (2.0) | 7.0 (2.0) |  |
| **Collective responsibility** | 5.6(1.6) | 6.7 (1.7) | 5.7 (1.7) | **P<0.001** |
| Q13: When everyone else is vaccinated, I don't have to be vaccinated, too. (R) | 5.0(3.0) | 7.0 (2.0) | 5.0 (3.0) |  |
| Q14: I get vaccinated because I can also protect people with a weaker immune system | 6.0(2.0) | 7.0 (1.0) | 6.0 (2.0) |  |
| Q15: Vaccination is a collective action to prevent the spread of diseases | 7.0(2.0) | 7.0 (1.0) | 7.0 (2.0) |  |

### Cutoff points determination

Table 3 and figure 1-5 showed the cutoff points of the different 5C sub-scales. The 5C questionnaire cutoff points were based on responses to the question’’ Did you get the COVID-19 vaccine?’’. One cutoff point was identified from ROC curve analysis for each of the 5C subscales. These cutoff points were used to classify respondents into:

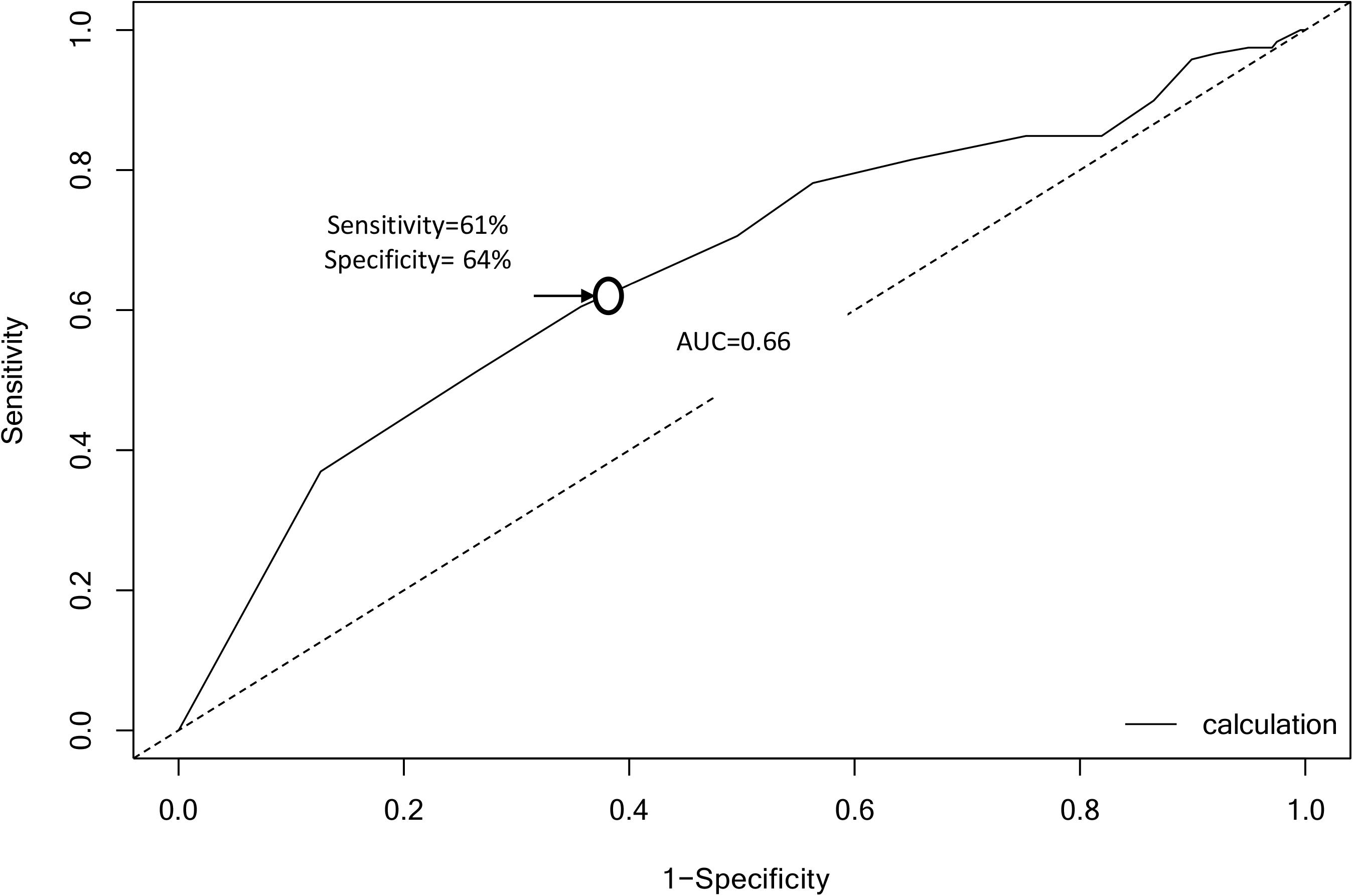

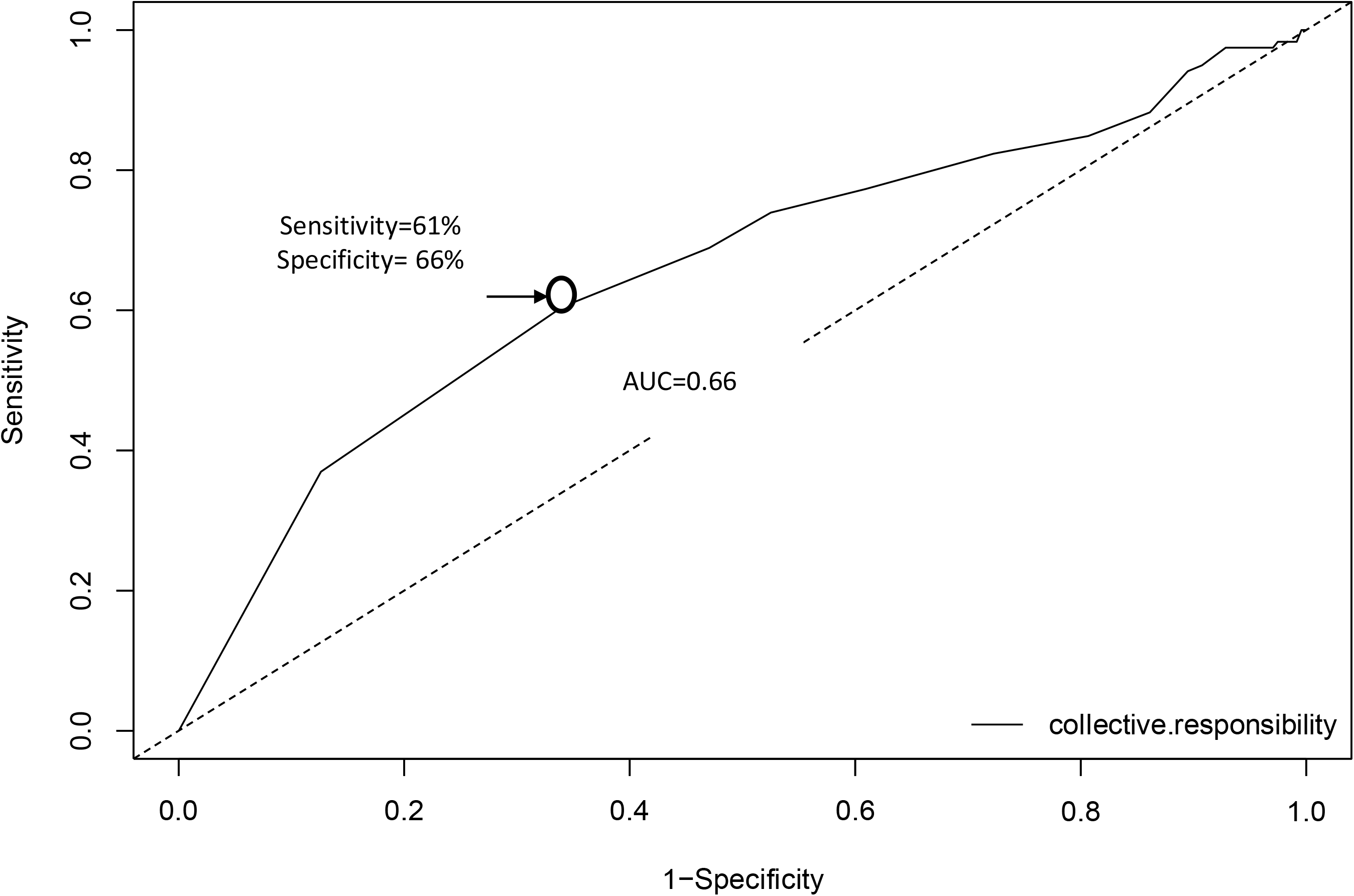

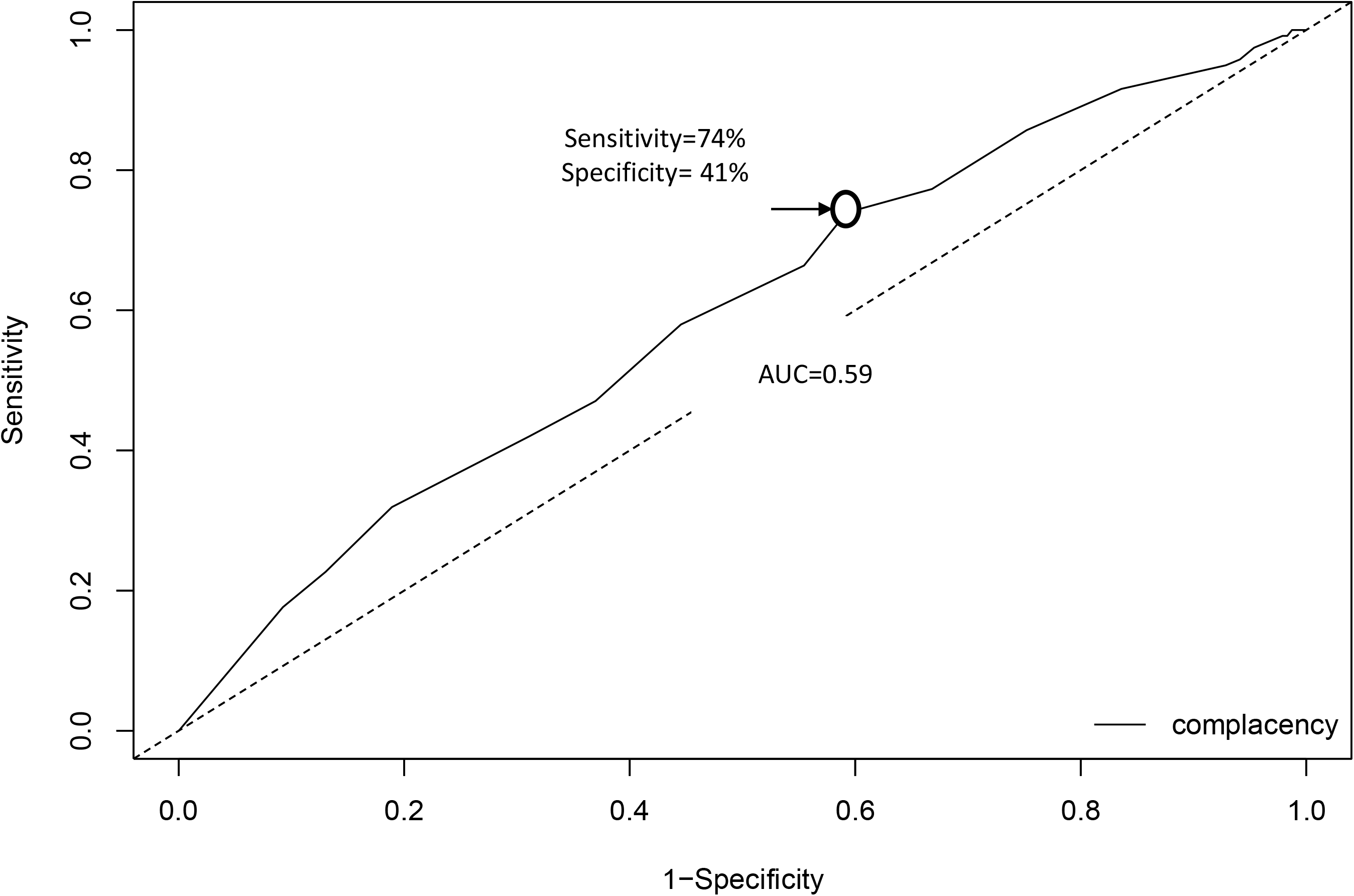

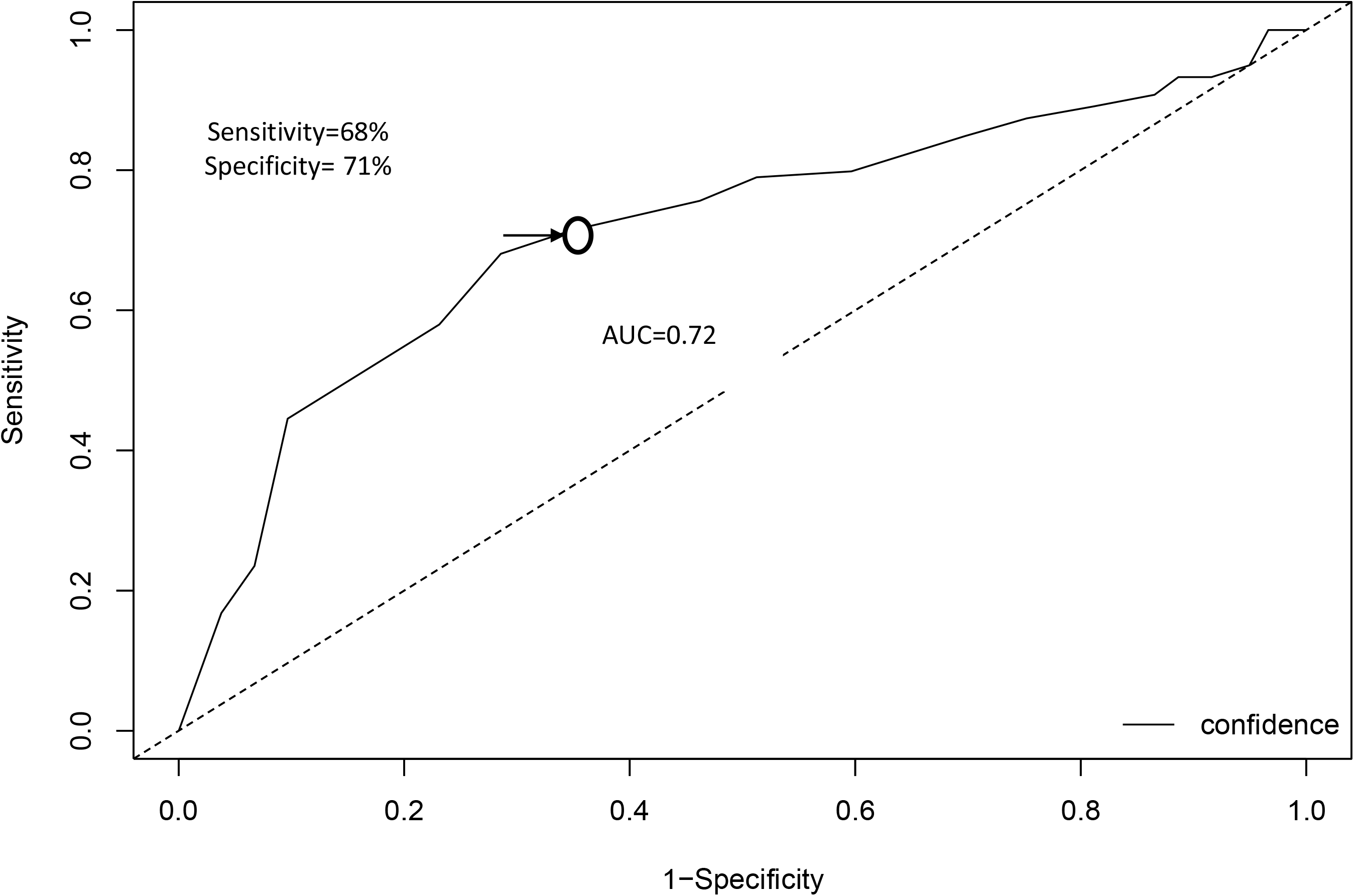

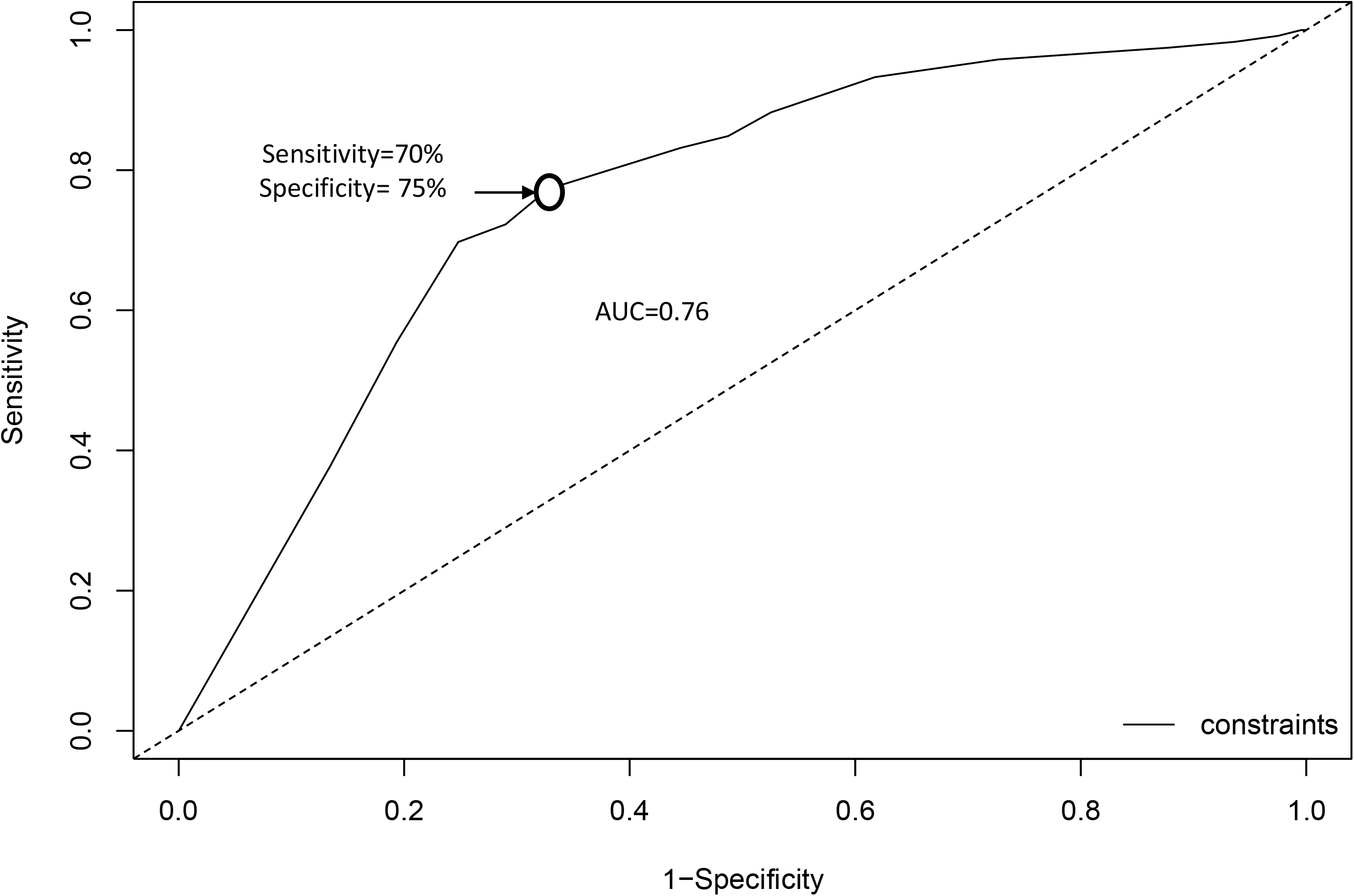

- Not confident: when confidence subscale score was <5.6, and confident when confidence subscale score was >= 5.6.
- Not complacent: when complacency score was < 4.7,and complacent when the subscale score was >=4.7.
- Did not have constraints against COVID-19 vaccine when the subscale score was <6.0, and had constraints when the sub-scale score was >=6.0.
- Did not assess different calculation for receiving COVID-19 vaccine when the sub-scale score was <6.3, and assessed calculation when the subscale score was >= 6.3
- Lacking (absence of) collective responsibility when the sub-score was <6.2, and having (feeling) collective responsibility when the subscale score was >=6.2.

**Table 3:** The cutoff values, sensitivity, and specificity of the 5C subscales.

| Subscale | Cutoff points | AUC (95% CI) | Sensitivity (95%CI) | Specificity (95%CI) |
| --- | --- | --- | --- | --- |
| **Confidence** | 5.7 | 0.72 (0.65- 0.78) | 0.68 (0.59-0.76) | 0.71 (0.65-0.77) |
| **Complacency** | 4.7 | 0.59 (0.53-0.66) | 0.74 (0.65-0.82) | 0.41 (0.35-0.47) |
| **Constraints** | 6.0 | 0.76 (0.71-0.81) | 0.70 (0.61-0.78) | 0.75 (0.69-0.81) |
| **Calculation** | 6.3 | 0.66 (0.60-0.72) | 0.61 (0.51-0.61) | 0.64 (0.58-0.73) |
| **Collective responsibility** | 6.2 | 0.66 (0.59-0.72) | 0.61 (0.51-0.69) | 0.66 (0.60-0.72) |

AUC: area under the curve, CI: confidence intervals

## Discussion

With the fast pace of COVID-19, researchers are becoming dependent on universal scales without evidence of effectiveness and efficacy within local population subgroups. This is critical as vaccine hesitancy, being a global public health issue, needs addressing for better controlling COVID-19 spread. This research was conducted to estimate a cutoff point at which each of the 5C domains can be assessed negatively or positively towards the COVID-19 vaccine’s attitude and behavior. Results indicated valuable cutoff points for which each domain could be utilized using proper test characteristics measures. As such, utilizing the Arabic version of the questionnaire is an asset as it discriminates against COVID-19 hesitancy utilizing a set of validated questions.

Developers of the 5C scale did not report clear cutoff points for its domains but rather recommended each implementer estimates cutoff point of each sub-scale according to the study context. (16) As it is clear from the stated definitions of the 5 subscales no umbrella concept embraces all antecedents. Thus, our results support the notion that calculating a uniform total score across all antecedents is not actually practical and that the sub-domains are of better use for presenting the dimensions of the scale. One of the major advantages of 5C scale is that it can assess the five antecedents is to explain vaccination behavior, to asses the need for interventional programs. If there is researcher discovered vaccine hesitany, the role of intervention come up to improve vaccine acceptanc.(19)

It is worthy noting that one of the benefits of the 5C scale is its ability to clearly detect early warning signs of vaccine hesitancy, consequently, guide policymakers and stakeholders to provide evidence-based public health interventional programs. The scale can be considered as an effective tool in the fight against COVID-19 consideing that effective non-pharmaceutical interventions are creating serious burdens on mental health status at the global level. Assessing and addressing vaccine hesitancy using a clear and tested cutoff point for 5C among Arab population subgroups is necessary and, given our results, more appropriate. The testing characteristics reported in the current study will be useful for future public health surveys that measure and explore COVID-19 vaccine hesitancy. Determining the cutoff point will be highly useful in identifying populations with higher hesitancy, hence, interventions could be easily directed towards them. Moreover, the result of this study will help in directing resources towards clear targets allowing for more effective vaccine hesitancy interventions.

In this study, vaccinated individuals reported higher education level, a higher number of working as health care professionals, receiving previous influenza vaccination at higher levels, fewer numbers of those previously infected with COVID-19, a higher proportion of relatives having previously infected with COVID-19 but less proportion of relatives dying due to COVID-19, and better awareness of different COVID-19 vaccines with higher percentage searching the web for information about COVID-19 vaccines. However, it was noticed that a higher proportion of those who reported that they were not vaccinated had at least one chronic disease. We speculate that this category’s refusal or hesitancy to get vaccinated may be due to the fear of the vaccine side effects that may extend beyond his fear from the infection itself and, possibly, due to the poor awareness about the need to get vaccinated being a vulnerable group. Another issue to be noticed was that a higher proportion of vaccinated participants have had relatives who were infected with COVID-19 even after being vaccinated. Although there was a higher presentation of health care workers among the vaccinated participants, we expect that they may receive the vaccine in the near future even after their colleagues and friends got the infection based on the WHO, Centers for Diseases Prevention and Control (CDC) and local public health department’s recommendations.(20)

Responses to the 5C scale determine what respondents think and feel about COVID-19 vaccines and being vaccinated. In this work, the cutoff point of the confidence scale was estimated at 5.7. Indicating that if the median score of the three items of this domain exceeded 5.7, then the respondent is more likely to be trusting the efficacy and safety of the COVID-19 vaccine, the system of vaccine delivery, and the policymakers. These cutoff points had a sensitivity and specificity of 68% and 71%, respectively.

For the complacency domain, which had a cutoff point of 4.7, the achieved sensitivity and specificity were reported at 74% and 40%, respectively. Indicating that if subjects scored above this value, then he or she probably thinks that vaccine is not necessary to prevent COVID-19 infection. In fact, a higher complacency score donates a lower perceived risk of COVID-19 diseases. Risk perception is a critical point to define when dealing with COVID-19 vaccination and for the development of proper communication messages that engage the general population against improper risk perception.

For constraints, a score at or above 6.0 was reported to have 70% sensitivity and 75% specificity in identifying lower constraints. Higher constraints mean poor access to health services, as well as a weak perception of self-efficacy and behavioral control. Thus, structural and psychological barriers (access, a lack of self-control) are “gate-keepers” against impeding the implementation of vaccination intentions into behavior. Travel time or inconvenient procedures may also act as barriers. Perceiving constraints should therefore be related to a lack of perceived behavioral control.(21)

For calculation, the fourth scale of the 5C, the area under the curve (0.66), and the median score (6.3) yielded 61% sensitivity and 64% specificity. Individuals with a score above this value have more perceived risk of COVID-19 infection as this scale donates that individuals weighted the risk of infection to that of vaccination. Depending on the information sources that are used, a high calculation can lead to non-vaccination due to the high availability of anti-vaccination sources, for instance, on the internet. (22) COVID-19 infodemic, therefore, could play a significant role in this item. Information shared over social media platforms is critical in defining this domain and should be considered when responses are higher than expected.

The last scale was a collective responsibility, with a median score of 6.2 or above has a sensitivity, and specificity of 61% and 66%, respectively, in determining the respondent behavior toward protecting others through self-vaccination. The respondent feels that he can protect others from contracting infection when they become vaccinated as it enhances herd immunity. A higher collective responsibility score usually correlates with empathy and collectivism of the respondents, while a lower score donates that the respondent doesn’t care about others and doesn’t know enough about herd immunity.(23) Social responsibility is a critical point to consider when dealing with COVID-19 vaccination hesitancy as it shows how responsible each individual to his community, the elderly, and those at higher risk. This domain deals with protective factors at the community levels as it may reflect how people behave during social gatherings.

## Data Availability

data available upon request by mailing the first author

## Limitation and strength

To the best of our knowledge, this is the first study in the Arab world that identifies a threshold for the 5C scale. Due to time restrictions, this survey was conducted only in three different Arab countries where vaccine delivery already started. We believe that conducting the study in three different countries is actually a strength as this may ensure the external validity of the research outcome. One of the major limitations is that this survey was being conducted online. An online survey has a lot of restrictions and limitations, but due to the national lockdown and national regulations, this was the most suitable method of data collection. The sampling technique was a non-random method, again due to the COVID-19 pandemic we could not select respondents randomly. One of the strength of this research is that the cutoff point determined would help researchers in the Arab world to use our previously validated 5C questionnaire and interpret population responses

### Data availability

Data are available upon request by mailing the first author

### Funding

This research was not funded.

### Author contribution

- **Ramy Mohamed Ghazy, Samar Abd ElHafeez, and Ramy Shaaban**: conceptualized the study idea, designed the survey tools, assisted in data collection, and composed the initial manuscript draft.
- **Ahmed Ramadan**: performed the statistical analysis, data interpretation, and visualization and participated in manuscript writing up.
- **Khalid Kheirallah:** assisted in data collection, interpretation, and visualization, edited the initial manuscript.
- **Iffat Elbarazi:** assisted in data collection, reviewing, and editing the manuscript
- **Marwa Shawky:** assisted in data collection and reviewing the manuscript.

